# Holistic antenatal care during pregnancy: A systematic review

**DOI:** 10.1101/2024.04.30.24306612

**Authors:** Violet Manonyana Chewe, Johanna Mmabojalwa Mathibe-Neke

## Abstract

**Background:** Pregnancy outcomes are determined by the type of antenatal care a woman receives. The purpose of this review was to conduct a critical analysis of the literature on the provision of holistic antenatal care to pregnant women.

**Design:** A systematic research review was employed for the study.

**Data Sources:** A search of EBSCO Host, Google Scholar, Sabinet, and Science Direct databases was performed. After reviewing 150 paper titles and abstracts, 14 studies were found to meet the final criteria for quality assessment. Included were only English-language papers published from 2020 to 2023.

**Eligibility Criteria for Selecting Studies:** Quantitative, Qualitative, Review studies, Quasi-experimental, and cohort studies. Papers had to include adults (>18 years old), and pregnant women attending antenatal care at different health facilities worldwide and published in peer-reviewed journals from 2020 onwards.

**Results:** Four of the fourteen analysed articles focused on socioeconomic status, three on staff attitudes, age, parity, and religion, and one on infrastructure. Six studies focused on midwives ’role in providing holistic antenatal care. These factors impacted pregnant women’s ability to receive comprehensive antenatal care. Three studies on woman-centered care and three on focused antenatal care have also come to light to enhance midwives’ capacity to deliver comprehensive prenatal care.

**Conclusion:** Healthcare facilities should offer an environment where communities may learn about the importance of antenatal care visits and how to ensure pregnant women obtain acceptable treatments.

**Implications:** This study demonstrates the necessity for healthcare providers to be resourceful about offering holistic antenatal care to pregnant women to enhance outcomes for both mothers and newborns.

## Introduction

Holistic antenatal care is the level of excellence in providing antenatal care to pregnant women to lower maternal mortality [1]. Antenatal care (ANC) is the care given to pregnant women by qualified healthcare professionals to provide the best possible health outcomes for both mother and child throughout pregnancy [2].

Other countries like the Netherlands have holistic midwives who offer integrated ANC to pregnant women [3]. A holistic midwife is a single midwife who interacts with a woman’s physical, psychological, and emotional factors during her pregnancy and birthing [4]. Moreover, [3] proposes that holistic midwives be utilized for high-quality ANC due to their inclusive services. The services include good relationships with women, mutual respect, and trust.

According to the findings from the study conducted in Bandung, Indonesia, by [5], holistic antenatal care lowers maternal anxiety levels in pregnant women. [5] further asserts that holistic ANC provides pregnant women with comprehensive psychological, emotional, social, and spiritual care. According to [6], pregnant women use antenatal care services for quality, which can be attained when provided holistically. On the other hand, [7] states that providing holistic ANC in particular situations can be challenging as some women attend ANC appointments solely to reserve a birthing location. Therefore, in such cases, women do not understand why it is crucial to attend ANC; hence, they end up skipping their scheduled follow-up appointments.

In a remote Tanzanian district, the quality of antenatal care was evaluated from a holistic angle [8]. The results showed that regular ANC services function inconsistently, partially accounted for by insufficient resources. Taking an acceptable history, paying attention to the client’s well-being, doing a basic physical examination, and providing proper counseling and education all displayed poor performance. Embracing holistic antenatal care services may benefit pregnant women and their unborn children.

As of the 2021 report, the Maternal Mortality Ratio (MMR) stood at 148.1 maternal deaths per 100,000 live births [9]. This is a 47% increase in maternal mortalities from 2019 and a further increase from the MMR of 126.1 in 2020. This demonstrates that in 2021 and 2020, the sharp fall between 2010 and 2019 has reversed. There was an increase in MMR from 2019 in every province in South Africa.

The purpose of this review was to identify patterns and conduct a critical analysis of the literature about the provision of holistic antenatal care. While there are many studies regarding antenatal care, this study focuses mainly on holistic antenatal care. Pregnant women should receive holistic ANC, which includes care for their physical, emotional, and spiritual needs [9].

Research question: What factors impact the provision of holistic antenatal care?

## Materials and Methods

A systematic review evaluated the factors that impact providing holistic antenatal care to pregnant women. There is no registration for the systematic review protocol.

### Eligibility Criteria

#### Inclusion

Studies were selected for the systematic review if they included adults (>18 years old) who attended antenatal care at public health facilities worldwide. Quantitative, Qualitative, Review, Quasi-experimental, and cohort studies were included. Studies had to have been published in peer-reviewed journals from 2020 onwards. Only articles published in English were considered for inclusion in the review due to the need for more resources available for translation services.

#### Exclusion

Grey literature, including conference abstracts, posters, and case reports, were excluded to ensure that only high-quality, peer-reviewed data was used in this systematic review. Published protocols with results were considered.

### Information sources

A subject librarian from the University of South Africa (UNISA) assisted with this process. The researcher used the following search engines to locate appropriate information for the study: EBSCO Host, Google Scholar, Sabinet, and Science Direct on 29 October 2023. The reference lists of non-experimental studies discovered through the electronic search and considered relevant to the review topic were also examined to identify relevant articles for inclusion in the next abstract review phase. There was no need for ethics approval because this was a review study.

The following attributes of each study in this review were extracted from the abstracts of the publications: author and year of publication, objective/aim, country, study design, population and sample size, outcomes, and limitations. Refer to Table 2 for the characteristics of the included studies.

#### Search Strategy

The researchers collaborated with the subject librarian to conduct a thorough literature search on all major electronic databases, including EBSCO Host, Google Scholar, Sabinet, and Science Direct. The studies extracted were those published between 2020 and 2024. The search parameters were antenatal care, holistic, care, and pregnancy to include all possibly relevant papers. The electronic databases used are listed in Table 1. The references of any non-experimental papers included in the search result were screened for relevant experimental studies.

**Table 1:** Electronic databases searched and the number of results.

| DATABASE | PAPERS IDENTIFIED |
| --- | --- |
| EBSCO Host, | 88 |
| Google Scholar, | 07 |
| Sabinet, | 40 |
| Science Direct | 15 |
| **Total** | **150** |

#### Selection process

Both authors VM and JM independently read the title and abstract of each paper to identify studies selected for full-text review based on the criteria mentioned above. Experimental studies whose interventions, patient populations, or outcomes could have been clearer based on the abstract, or papers that did not have an abstract available, were also moved on to full-text review for further assessment. Full texts of papers were also screened for mentions of the “Holistic/ quality antenatal care, midwives, pregnant women, quality antenatal care” population for the study. Papers with no references to these words were considered not to include populations of interest and were excluded from the full-text stage of the review. Finally, eligible papers deemed to meet inclusion and exclusion criteria following the full-text review underwent quality appraisal and final inclusion in the systematic review. Any discrepancies were resolved by both the author and the co-author.

The researcher outlined the study flow in the search results using the PRISMA flow chart suggested by [10] (see Figure 1). The search yielded a total of 150 articles. In these papers, the researchers conducted full-text, topic, and abstract screenings; as a result, duplicates were eliminated, and studies that didn’t meet the inclusion criteria were disqualified. Ultimately, it was concluded that 14 papers were appropriate for inclusion in the analysis since they addressed the study’s purpose, focus, and review question.

**Fig 1:**
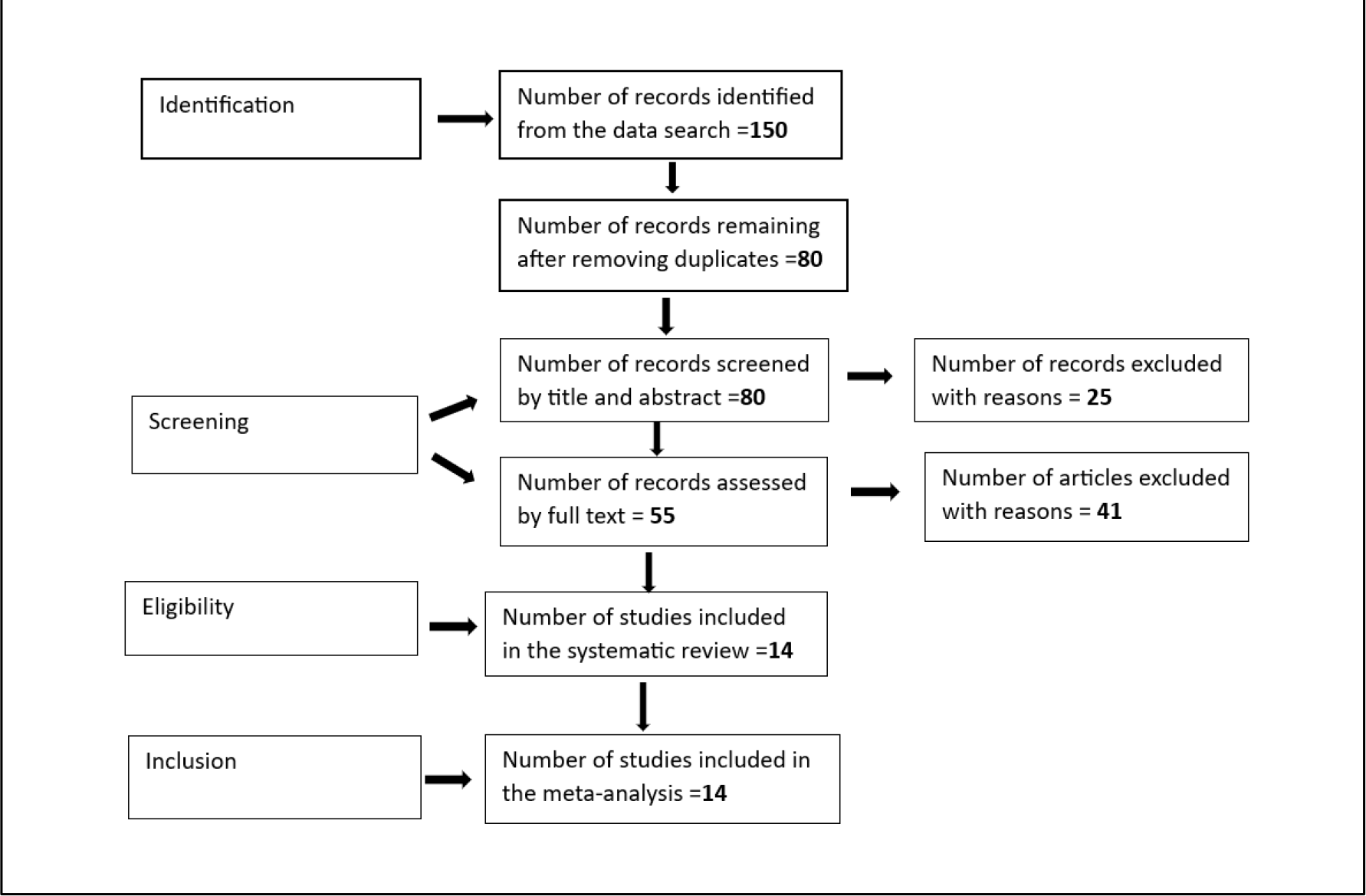
Screening process flow chart

#### Data collection process

The author and the co-author independently extracted relevant information from each paper. Where information still needed to be included after reviewing those sources, the papers were excluded from the review.

#### Data Items

Specific study-related outcomes were extracted from each paper by applying the data from the original publication. Data on the study demographic, participant number, holistic/high-quality prenatal care, and outcomes reported in the original study were collected from each publication to create Table 2.

**Table 2:** Characteristics of the studies included.

| Author and year | Aim/Objective | Country | Study design | Population and sample size | Outcomes | Limitations |
| --- | --- | --- | --- | --- | --- | --- |
| Yehualash | To | Ethiopia | Secondary | 2019 | A woman's place of | A |
| et et al<br>2021, | determine<br>the factors<br>related to<br>the best<br>ANC visits<br>for<br>Ethiopian<br>pregnant<br>women |  | data<br>analysis | Ethiopian<br>Mini<br>Demograp<br>hic and<br>Health<br>Survey<br>(EMDHS | residence, financial<br>status, media<br>exposure,<br>education level,<br>and geography are<br>all closely linked<br>with a suitable<br>timing for an ANC<br>visit. | community's<br>and an<br>individual's<br>level factor<br>that<br>contribute to<br>the ideal<br>number of<br>ANC visits<br>aren't<br>demonstrate<br>d by the<br>cross-sectional<br>data. |
| Andegiorgi<br>sh et al<br>2022, | To explore<br>the variables<br>related to<br>antenatal<br>care (ANC)<br>visits | SSA | Secondary<br>data s<br>analysis<br>using<br>cross-sectional<br>research<br>data. | 56<br>002wome<br>n aged 15–<br>49 years in<br>Ghana<br>(3224),<br>Kenya (10<br>981),<br>Malawi<br>(9541),<br>Namibia<br>(2286),<br>Rwanda<br>(4416),<br>Senegal<br>(6552),<br>Tanzania<br>(5536),<br>Uganda<br>(7979) and<br>Zambia | There was a<br>significant<br>difference in the<br>number of ANC<br>visits among the<br>countries under<br>study, with the<br>amount deemed<br>inadequate. | Not stated |
|  |  |  |  | (5487)<br>were<br>analysed |  |  |
| Kare et al<br>2021 | The purpose<br>of the<br>research<br>was to<br>evaluate the<br>standard of<br>ANC and<br>related<br>factors for<br>pregnant<br>women who<br>were<br>admitted to<br>government<br>institutions | Ethiopia | Cross-sectional<br>study<br>(Quantitati<br>ve) | 372<br>pregnant<br>women. | The number of<br>antenatal visits,<br>residency, financial<br>status, and<br>educational<br>attainment were all<br>linked to high-quality healthcare. | As the study<br>was<br>conducted at<br>a hospital,<br>probably, the<br>results<br>cannot be<br>applied to all<br>pregnant<br>women in<br>the area. |
| Ahinkorah<br>et al 2021. | This study<br>examined<br>the<br>relationship<br>between<br>pregnant<br>women<br>seeking<br>prenatal<br>care in a<br>few sub-Saharan<br>African<br>countries<br>and the<br>implementat<br>ion of the<br>2016 WHO antenatal care services model. | SSA | The study<br>solely<br>considered<br>data from<br>the 2018<br>Sub-Saharan<br>African<br>Demograp<br>hic and<br>Health<br>Survey. | 6761 from<br>Nigeria,<br>1973 from<br>Mali, 1690<br>from<br>Guinea,<br>and 1570<br>from<br>Zambia<br>were<br>considered<br>. | The study<br>demonstrated the<br>impact that<br>healthcare access<br>restrictions have on<br>the use of antenatal<br>care in sub-Saharan African<br>nations. | Most of the<br>queries came<br>from married<br>women;<br>hence, only<br>married<br>women were<br>qualified. |
| Appiah 2022 | The study investigated the connections between individual and community factors and early ANC use in Cameroon. | Cameroon | A cross-sectional survey design | 4,183 women | Early ANC initiation correlated with individual-level factors (maternal age, economic status, academic success, and religious affiliation), while variations in cluster/community depict influenced early ANC initiation. | In this study, only the association between early ANC utilization and individual and community variables was evaluated |
| Warri and George 2020. | The study aimed to determine why pregnant women at Nkwen Baptist Health Center delayed starting prenatal care. | Cameroon | Exploratory, qualitative study | Eighteen pregnant women | The time of initiating antenatal care is significantly influenced by sociocultural beliefs and barriers found in health facilities. | Not stated |
| Seyoum 2022. | The study aimed to identify the factors | Ethiopia | Cross-Sectional Studies. | 13 studies | Determinants of antenatal care service satisfaction were the first | One limitation of this analysis was the |
|  | that affect Ethiopian women's satisfaction with antenatal services. |  |  |  | antenatal visit, waiting time to see the practitioner, privacy preservation, courteous treatment, and unplanned pregnancy. | significant differences across the included research. |
| Hlongwane et al 2021. | The study described the implementation phases and the key indicator monitoring process. | SA | An analysis was conducted on samples of maternity case records from the first year of the modified care model | 200 pregnant women | The experiences of South Africa with the modified antenatal care package execution demonstrate that success requires the dedication of all parties involved. | Not stated |
| Miller et al 2021. | The study investigated the prevalence of PCANC in Rwanda and its contributing factors. | Rwanda | Quantitative data | 2150 surveys | The implementation of group care, which would completely restructure the way ANC is delivered, could interfere with delivering essential care elements, particularly those that are person- centered. | was not the study's intention to quantify PCANC specifically. |
| Swift et al 2021. | The purpose of the study was to evaluate EAC in terms of women's autonomy in making decisions, their level of happiness with care, and how well it reduces their dread of birthing. | Australia | Quasi-experimental controlled trial | 32 nulliparous women, 60 usual antenatal care = 92 | In general, EAC seemed to be more successful than standard care in lowering birth anxiety, particularly in women who did not attend workshops in addition to prenatal care. | Small and homogenous sample size |
| Nagineviciute et al 2023. | The study employs patient-reported outcome and experience measures to assess low-risk pregnancies in Lithuania that are cared for by midwives and obstetrician- gynaecologists | Europe | A prospective cohort study design | 153 Pregnant women | Healthcare systems can better understand and respond to women's needs and preferences by using patient-centered approaches, which promotes high-quality, woman-centered care. | comparatively small sample size |
| Konlan et al 2020 | The purpose of the study was to evaluate the factors that affect pregnant women's utilization of FANC services. | Ghana | Cross-sectional quantitative study | 210 postnatal women | Some postnatal women are unaware of FANC, even though it would assist the health professional in providing information, education, and communication tools, as the results showed. | Not stated |
| Nwabueze et al 2023, | The purpose of this study was to evaluate pregnant women in Nigeria's knowledge and use of FANC, as well as the factors that affect their use of it. | Nigeria | A cross-sectional design | 300 pregnant women | The results of this study hold significant ramifications for formulating interventions and policies related to maternal and child health that will enhance Nigerian women's and children's access to healthcare throughout pregnancy and delivery. | The study was limited to pregnant patients who receive care at public institutions. |
| Abera et al 2020. | This study sought to evaluate the parameters related to focused | Ethiopia | Cross-sectional study | 551 women who gave birth in the last two years | Improve programs that encourage pregnancy planning, shorten travel times to medical facilities, | Not stated |
|  | antenatal care (FANC) attendance in Wolaita Zone, Southern Ethiopia, and the utilization of these services. |  |  |  | and raise pregnant women's knowledge of the advantages of antenatal care. |  |

#### Study risk of bias assessment

The Revised Cochrane risk-of-bias tool for randomized trials (RoB 2) was used for randomized, controlled trials in the final publication batch [11]. VM and JM evaluated the tool’s signaling questions individually to evaluate each paper’s quality and bias risk. After the meeting, VM and JM agreed on any differences in their assessments of the quality of the work. Similarly, the overall risk of bias in the quasi-experimental study was calculated using Tables 1 and 2 of the ROBINS-I tool [12]. VM and JM carried out the quality assessment in the process stated above.

## Results

The analysis approach of choice was thematic, employing Caulfield’s method [13]. It aided the researchers’ familiarisation with the data as they read the articles and abstracts. After that, during the coding process, the researchers highlighted terms, phrases, and conclusions that often emerged in the data. The researchers established themes from these, as shown in Table 3, which led to the findings and discussions.

**Table 3:** Themes and sub-themes.

| THEMES | SUB-THEMES | ARTICLES |
| --- | --- | --- |
| 1. Factors affecting the provision of quality antenatal | 1.1 Socio-Economic factors | ➤ Yehualashet, Seboka, Tesfa, Mamo, and Seid 2021 |
| care. |  | ➤ Andegiorgish, Elhoumed, Qi, Zhu, and Zeng 2022,<br>➤ Kare, Gujo & Yote 2021<br>➤ Ahinkorah, Ameyaw, Seidu, Odusina, Keetile, and Yaya 2021 |
|  | 1.2 Marital status, age, parity, and religion | ➤ Appiah 2022<br>➤ Konlan, Saah, Amoah, Doat, Mohammed, Abdulai, & Konlan 2020<br>➤ Warri & George 2020 |
|  | 1.3 Staff attitude | ➤ Warri and George 2020,<br>➤ Seyoum 2022 |
|  | 1.4 Infrastructure | ➤ Hlongwane, Bozkurt, Barreix, Pattinson, Gülmezoglu, Vannevela & Tunçalp 2021. |
| 2. The role of midwives in providing holistic antenatal care | 2.1 Woman-Centred Approach | ➤ Miller, Afulani, Musange, Sayingoza, and Walker 2021<br>➤ Swift, Zoega, Stoll, Avery, and Gottfreðsdóttir 2021<br>➤ Nagineviciute, Bartuseviciene, & Blazeviciene 2023. |
|  | 2.2 Focused Antenatal Care | ➤ Konlan, Saah, Amoah, Doat, Mohammed, Abdulai, & Konlan 2020,<br>➤ Nwabueze, Okeke, Nwebo, Nwondo, Nwekpa & Nwaiwu 2023,<br>➤ Abera, Asseffa, Obssa, Balla, and Koyira 2020. |

## Discussion

Socioeconomic disparities in terms of ANC services impact certain localities. For example, financial and geographical barriers make it difficult for American doctors to provide women with the antenatal care they require [14]. The study by [15] claims wealthier women are advantaged as they can pay and access healthcare services. This demonstrates how income, education, and residence location can contribute to disparities in access to ANC services. According to [16], women living in rural Sub-Saharan nations are more likely to receive substandard ANC due to low incomes and a location of residence. The study by [17] found additional socioeconomic determinants, such as a husband’s acceptance. Despite the obstacles mentioned, Muslim pregnant women in Nigeria receive early ANC timing [17].

Maternal age, parity, and marital status appear to impact the use of ANC services in certain cases. [18] claims that young, single women are hesitant to seek antenatal care when pregnant. Additional research reveals that women with high parity are reluctant to use antenatal services due to a history of healthy pregnancies, lack of need, and other factors [19]. Quality ANC is additionally related to other factors, such as the woman’s behavioral and psychological well-being and the number of antenatal visits she had [1].

In most African countries, including South Africa, women hold a range of religious beliefs, which could influence how they use ANC services. According to [20, 17], pregnant women who experience stigma associated with their pregnancy, fear of witchcraft, cultural beliefs, and problems with family support are less likely to use antenatal services.

Healthcare personnel should always have a welcoming approach, according to one of the ministerial goals of South Africa [21]. According to [20], concerns were made about the staff members’ lack of courtesy. A further obstacle that has been identified as discouraging pregnant women from accessing ANC is the staff members’ disrespect towards them [22]. As such, midwives should welcome pregnant women positively and use the appointment system to commence making amends.

The results additionally indicated that pregnant women are more reluctant to be treated at healthcare facilities with inadequate, clean, or well-kept amenities. To provide pregnant women in SA with holistic ANC and lower the rate of maternal and newborn mortality, commitment and resource allocation are required [23]. Health facilities should regularly maintain their infrastructure, as it is mentioned as another factor that contributes to pregnant women’s poor attendance and inadequate ANC.

Woman-centered care prioritizes each woman’s needs, goals, and aspirations ahead of those of the institution or the midwifery profession. According to [24], women reported feeling disrespected and leaving ANC confused or with questions. Enhancing ANC was considered a means of improving the standard of ANC. According to a study conducted in 2021 by [25], women who received better ANC were more inclined than those who received traditional care. Furthermore, woman-centered approaches enable healthcare systems to identify and address the needs and preferences of women, encouraging the provision of high-quality ANC [26]. Given this, delivery anxiety was much reduced by improved ANC as compared to standard treatment.

The World Health Organization created focused antenatal care (FANC), a novel type of ANC, in 2001 with the goal of using it in underdeveloped countries [19]. While a pregnant woman participates in FANC, she attends at least four scheduled ANC visits. She receives all comprehensive packages from registered healthcare practitioners recommended by the World Health Organization [2]. The quality of services rather than their quantity matters under this strategy.

Although the overall ANC use might have seemed adequate, the strategy needed to be more comprehensive, targeted, and consistent with current norms. For example, [27] demonstrates how inadequately pregnant women use FANC. Furthermore, according to [28], there is little knowledge about the method in pregnant women. Thus, the emphasis of antenatal treatment continues to be on clinical evaluation and regular clinic visits, despite recognizing the importance of FANC.

## Strength and limitations

The authors VM and JM are experts in midwifery care who are currently practicing in academia, with antenatal care as their niche area of research. The analysis considered papers from international, Sub-Saharan African, African, and South African publications. The study was limited by current articles that matched the inclusion criteria.

## Conclusion

The systematic review identified issues affecting pregnant women’s access to holistic ANC. These include facility infrastructure, staff attitudes, age, parity, religion, and socioeconomic status. Nevertheless, other research acknowledged FANC and woman-centered care as ways to improve midwives’ ability to provide holistic ANC. It might make sense for healthcare institutions to provide a setting where residents are informed about the crucial nature of ANC consultations and how they can ensure that pregnant women obtain safe services.

## Data Availability

The author can give the data supporting the study's findings upon request.

## Acknowledgment

The researchers’ independent research from antenatal and maternal health research studies is presented in this article. Therefore, only writers’ opinions and points of view are expressed.

## Supporting information

S1 File. (ZIP)

## Author contribution

VM and JM were both responsible for conceptualization, data curation, methodology, analysis writing of the draft, review, and manuscript editing.

